# Clinical characteristics of 50466 patients with 2019-nCoV infection

**DOI:** 10.1101/2020.02.18.20024539

**Authors:** Pengfei Sun, Shuyan Qie, Zongjan Liu, Jizhen Ren, Jianing Xi

## Abstract

**Objective:** We aim to summarize reliable evidences of evidence-based medicine for the treatment and prevention of the 2019 novel coronavirus (2019-nCoV) by analyzing all the published studies on the clinical characteristics of patients with 2019-nCoV.

**Methods:** PubMed, Cochrane Library, Embase, and other databases were searched. Several studies on the clinical characteristics of 2019-nCoV infection were collected for Meta-analysis.

**Results:** Ten studies were included in Meta-analysis, including a total number of 50466 patients with 2019-nCoV infection. Meta-analysis shows that, among these patients, the incidence of fever was 89.1%, the incidence of cough was 72.2%, and the incidence of muscle soreness or fatigue was 42.5%. The incidence of acute respiratory distress syndrome (ARDS) was 14.8%, the incidence of abnormal chest computer tomography (CT) was 96.6%, the percentage of severe cases in all infected cases was 18.1%, and the case fatality rate of patients with 2019-nCoV infection was 4.3%.

**Conclusion:** Fever and cough are the most common symptoms in patients with 2019-nCoV infection, and most of these patients have abnormal chest CT examination. Several people have muscle soreness or fatigue as well as ARDS. Diarrhea, hemoptysis, headache, sore throat, shock, and other symptoms only occur in a small number of patients. The case fatality rate of patients with 2019-nCoV infection is lower than that of Severe Acute Respiratory Syndrome (SARS) and Middle East Respiratory Syndrome (MERS).

## 1 Introduction

Since December 2019, the epidemic of 2019 novel coronavirus (2019-nCoV) infectious pneumonia in Wuhan, China. The Chinese government and researchers have taken rapid measures to control the epidemic ^[1]^. On January 30, 2020, WHO declared that the epidemic of 2019-nCoV was a public health emergency of international concern (PHEIC). At present, the number of patients with 2019-nCoV infection is still rising, and its harm to human beings has exceeded the outbreak of Severe Acute Respiratory Syndrome (SRAS) in China, 2002 ^[2]^. The clinical characteristics and the case fatality rate after 2019-nCoV infection have always been concerned by people, and it is also the focus of medical workers’ research at present. However, due to the different design of different clinical studies and insufficient sample size, the conclusions of the published studies were different. In order to acquire more accurate conclusions on the clinical characteristics and mortality of patients with 2019-nCoV infection, we searched the relevant literatures and carried out single arm Meta-analysis^[3]^. Our findings provide important guidance for current clinical work on the prevention and treatment of 2019-nCoV infection.

## 2 Material and Methods

### 2.1 Search strategy

Three popular medical databases including PubMed, Cochrane Library, and Embase databases were searched for related literatures, using the following keywords: “2019-nCoV”, “Coronavirus”, “COVID-19”, “SARS-CoV-2” and “Wuhan Coronavirus”. Articles reviewed were dated up to February 2020. In this Meta-analysis, there was no language restriction. Only available data from published articles were collected. Data from unpublished papers were not included.

### 2.2 The inclusive and exclusive criteria

#### 2.2.1 Inclusive criteria

studies that include randomized controlled trials, non-randomized controlled trials, case-control studies, cohort studies, cross-sectional studies, and also case reports on the clinical characteristics of patients with 2019-nCoV infection. Clinical characteristics of patients with 2019-nCoV infection include fever, cough, muscle soreness or fatigue, acute respiratory distress syndrome (ARDS), abnormal chest computer tomography (CT) detection, patients in critical condition, as well as death due to 2019-nCoV infection.

#### 2.2.2 Exclusive criteria

articles were published repeatedly; studies did not include the research indicators needed for Meta-analysis; research data were missing.

### 2.3 Data extraction and paper quality evaluation

First of all, we screened the literature according to the literature abstract, excluding the articles that obviously do not meet the inclusive criteria, and then read the full article for re-screening. If any disagreement on the choice of the literature exists, a third evaluator will join to make the decision. All included literature were evaluated using the Newcastle-Ottawa Scale (NOS) ^[4]^. The highest quality of the literature was 9 stars and the lowest 0 stars.

### 2.4 Statistical analysis

The statistical software *Stata* version 12.0 was used to carry out the single arm Meta-analysis. Original data included in the literature were first transformed by double arcsine method to make them conform to normal distribution and then analyzed in *Stata*. The initial conclusion obtained by Meta-analysis was then restored using formula (P=(sin(tp/2))^2^) to reach final conclusion. In order to objectively evaluate the publication bias of the included literature, the Egger test with P < 0.05 as the existence of publication bias was performed, the values larger than which were considered as no publication bias.

## 3 Results

### 3.1 Literature inclusion

A total of 284 articles were retrieved, among which 39 papers were removed due to repeated retrieval, 212 papers were removed after reading abstracts, 23 were eliminated after reading the full text. At the end, a total of 10 articles ^[5-14]^ were included in this Meta-analysis, including data from 50466 patients. The specific operation flow is shown in Figure 1. The characteristics of the literature are shown in Table 1.

**Table 1:**
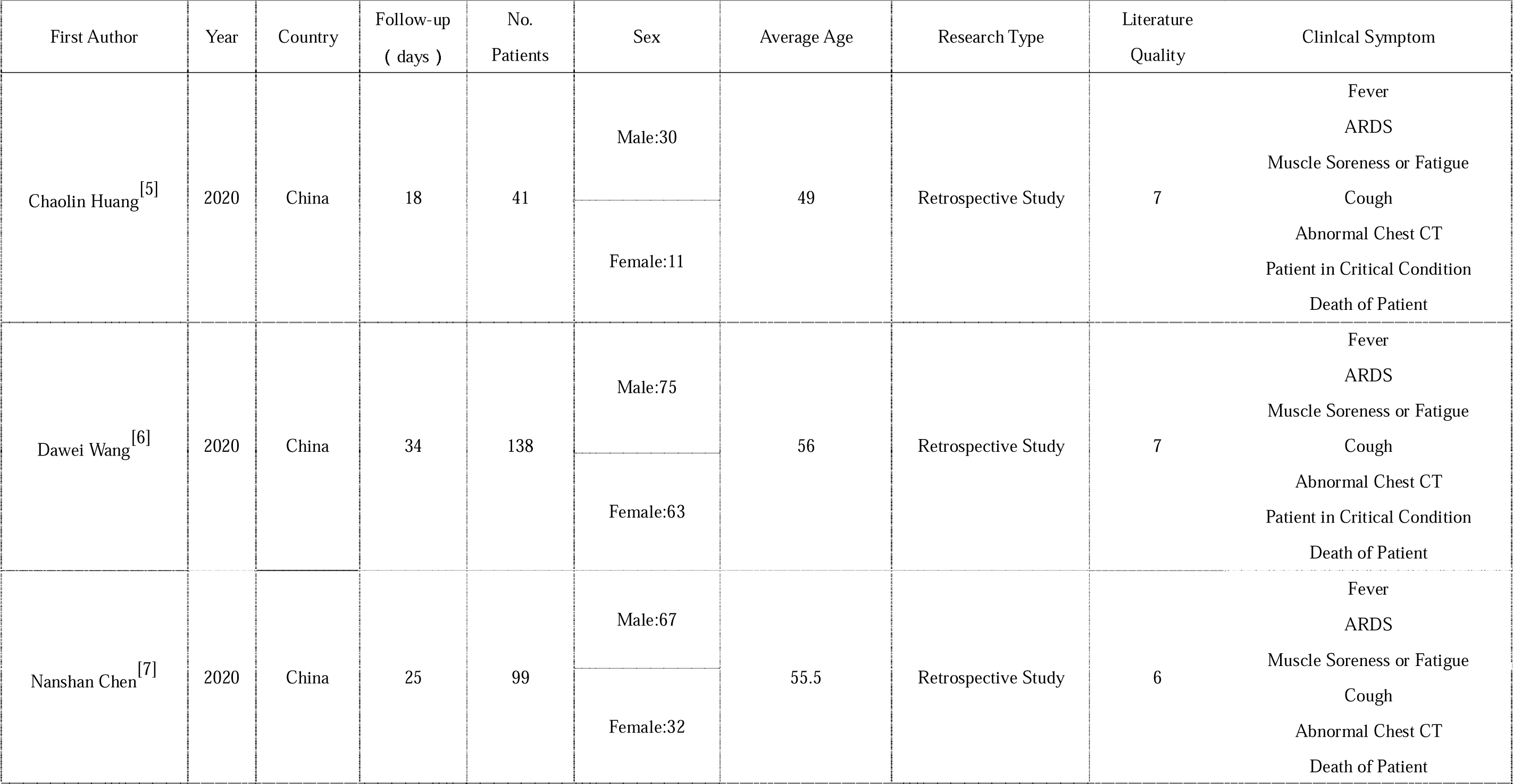
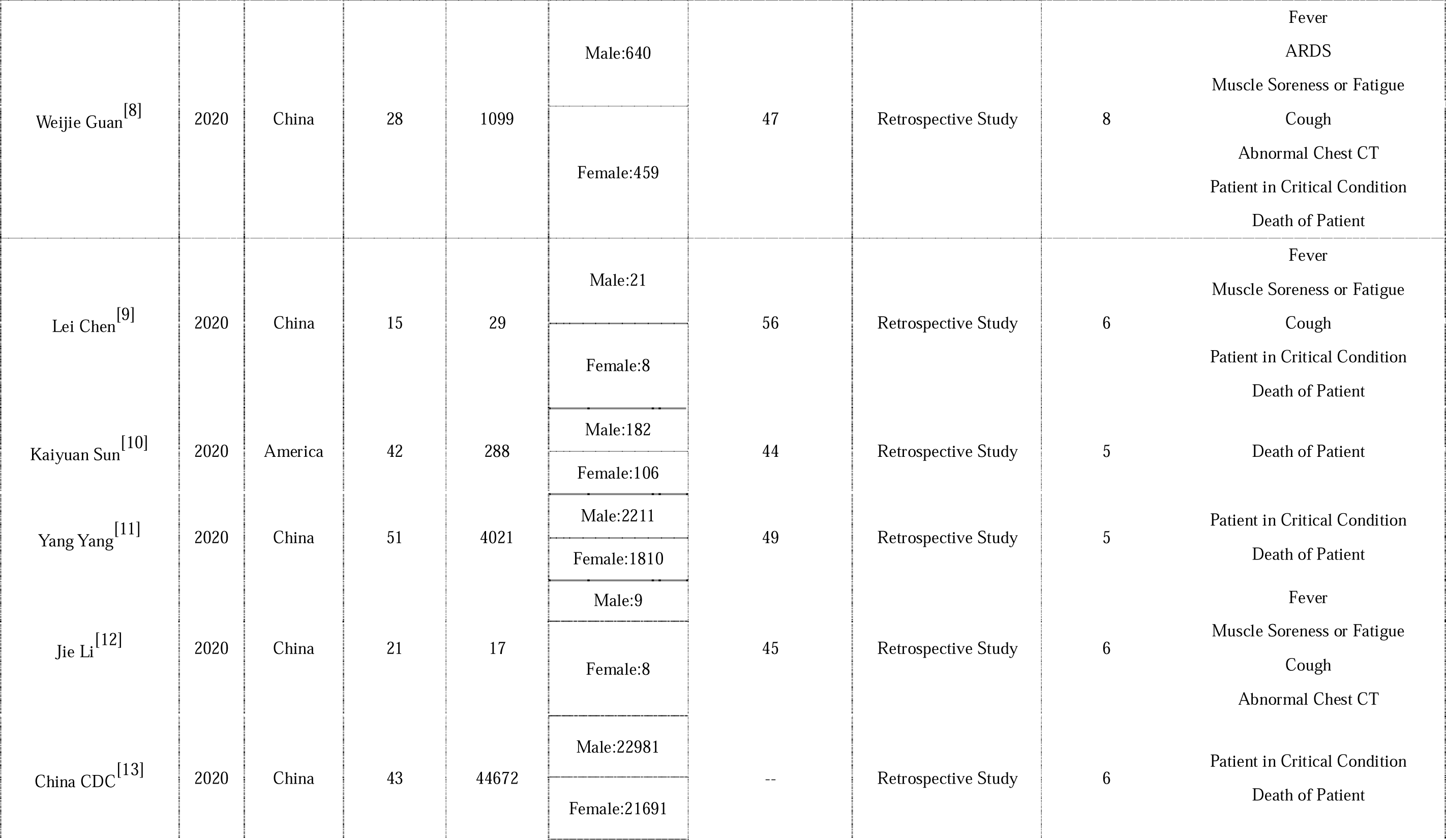
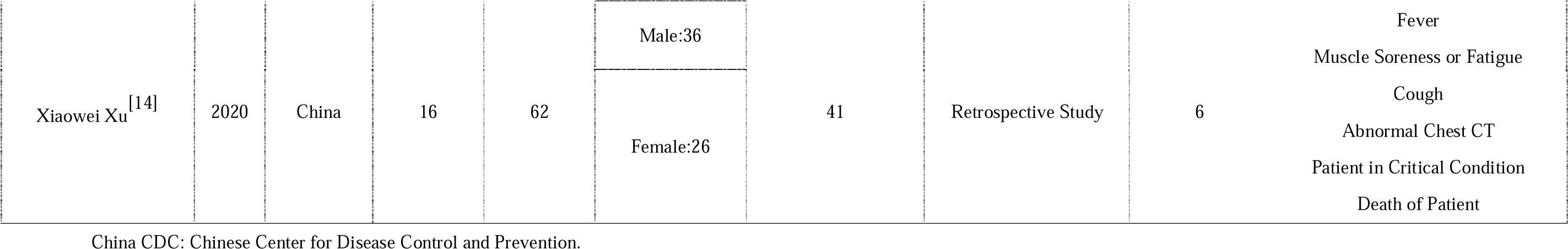
The characteristics of the literature.

**Figure 1.**
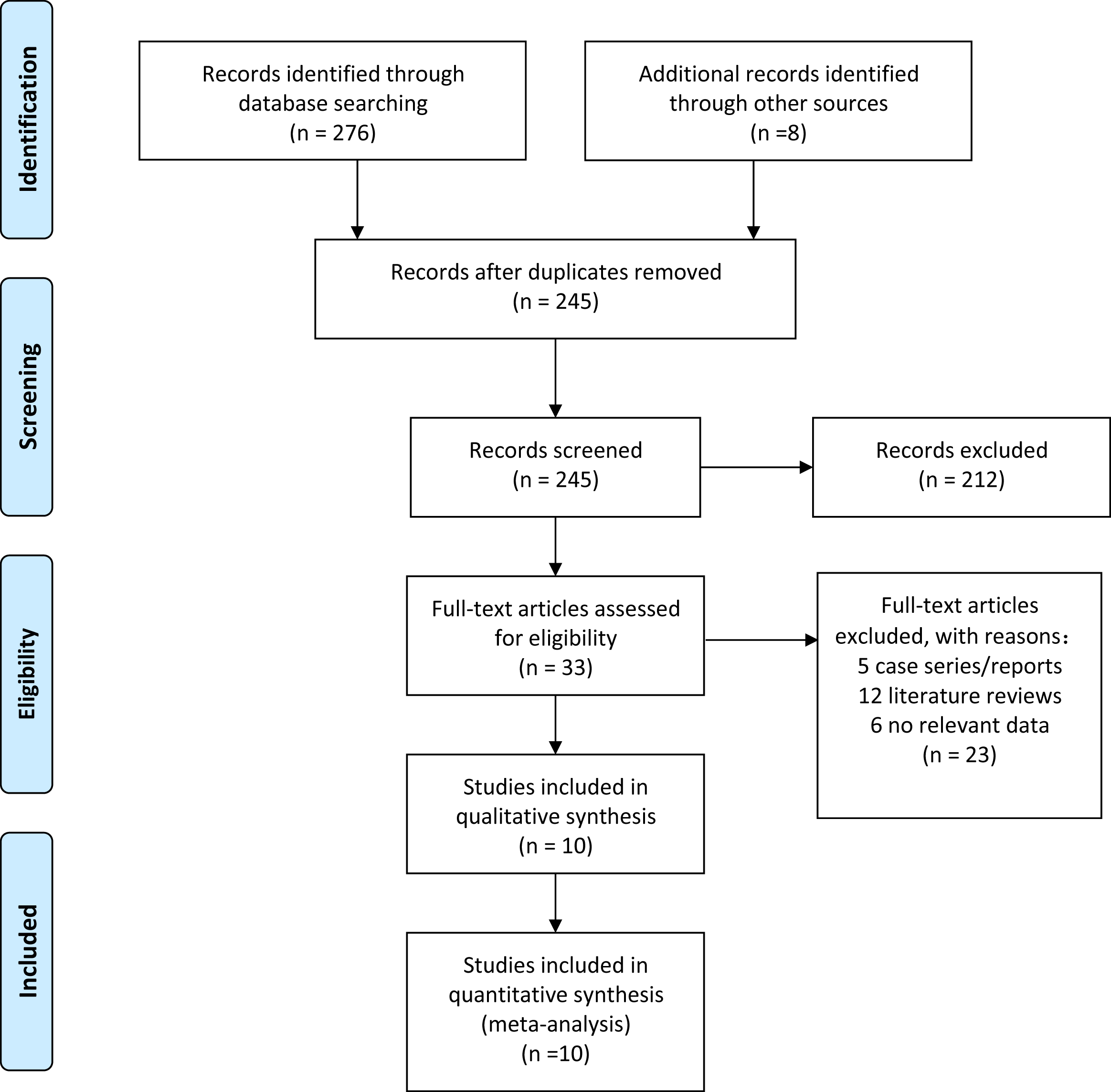
PRISMA flow diagram.

### 3.2 Meta-analysis results

Through Meta-analysis, we found that among all the clinical characteristics of patients with 2019-nCoV infection, the incidence of fever was 89.1% (Figure 2), that of cough was 72.2% (Figure 3), and that of muscle soreness or fatigue was 42.5% (Figure 4). The incidence of ARDS was 14.8% (Figure 5), correspondingly that of abnormal chest CT detection was 96.6% (Figure 6), severe cases in all infected cases occupied a percentage of 18.1% (Figure 7). The case fatality rate of patients with 2019-nCoV infection was 4.3% (Figure 8). Detailed results of Meta-analysis are shown in Table 2.

**Table 2:** Meta-analysis results.

| Clinlcal Symptom | Results of Meta-analysis | Adjusted Results |
| --- | --- | --- |
| Fever | 2.47(2.26,2.67) | 0.891(0.818,0.945) |
| ARDS | 0.79(0.43,1.15) | 0.148(0.046,0.296) |
| Muscle Soreness or Fatigue | 1.42(0.96,1.88) | 0.425(0.213,0.652) |
| Cough | 2.03(1.89,2.17) | 0.722(0.657,0.782) |
| Abnormal Chest CT | 2.77(2.57,2.97) | 0.966(0.921,0.993) |
| Patient in Critical Condition | 0.88(0.73,1.03) | 0.181(0.127,0.243) |
| Death of Patient | 0.42(0.33,0.50) | 0.043(0.027,0.061) |

**Figure 2.**
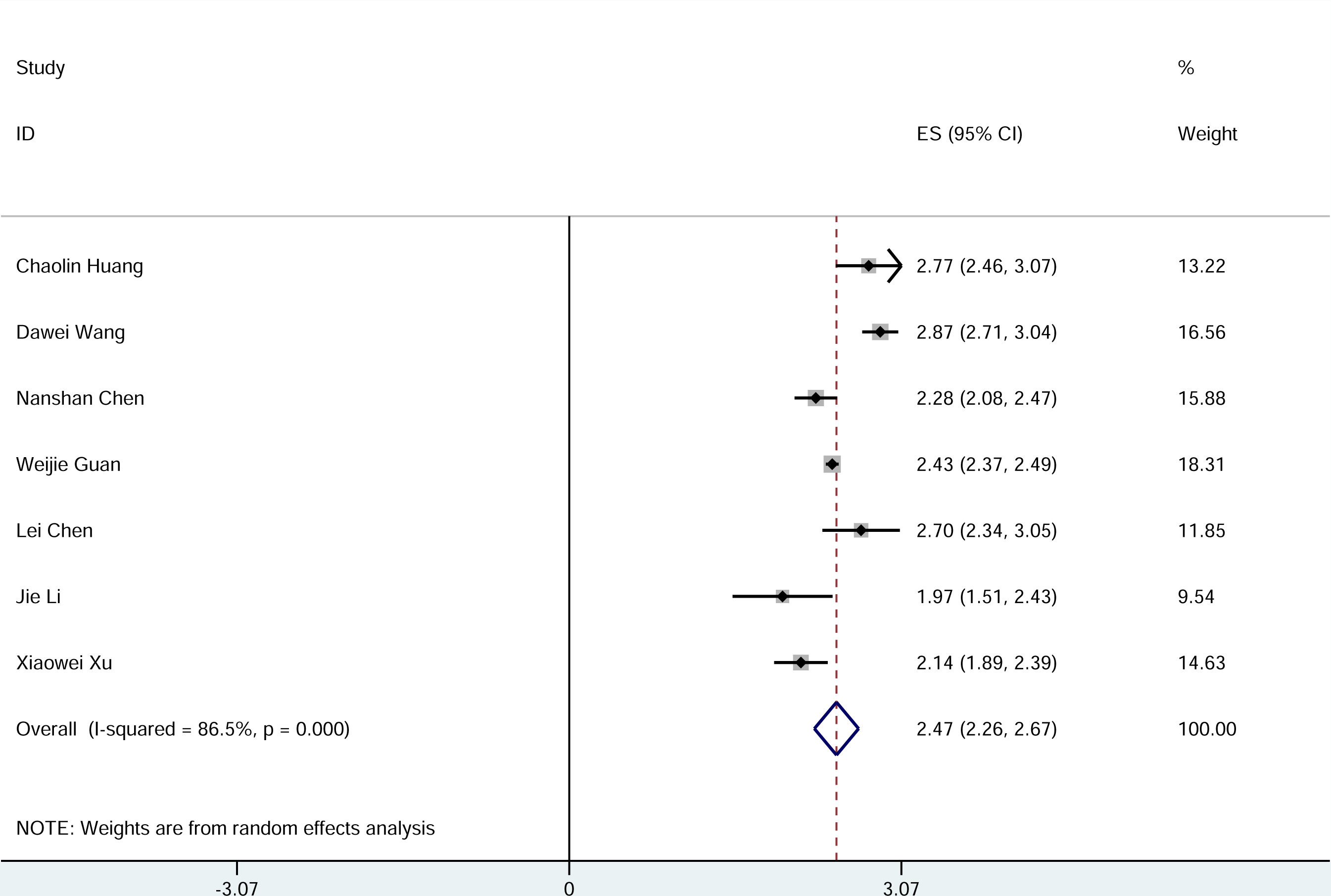
Meta-analysis of the incidence of fever.

**Figure 3.**
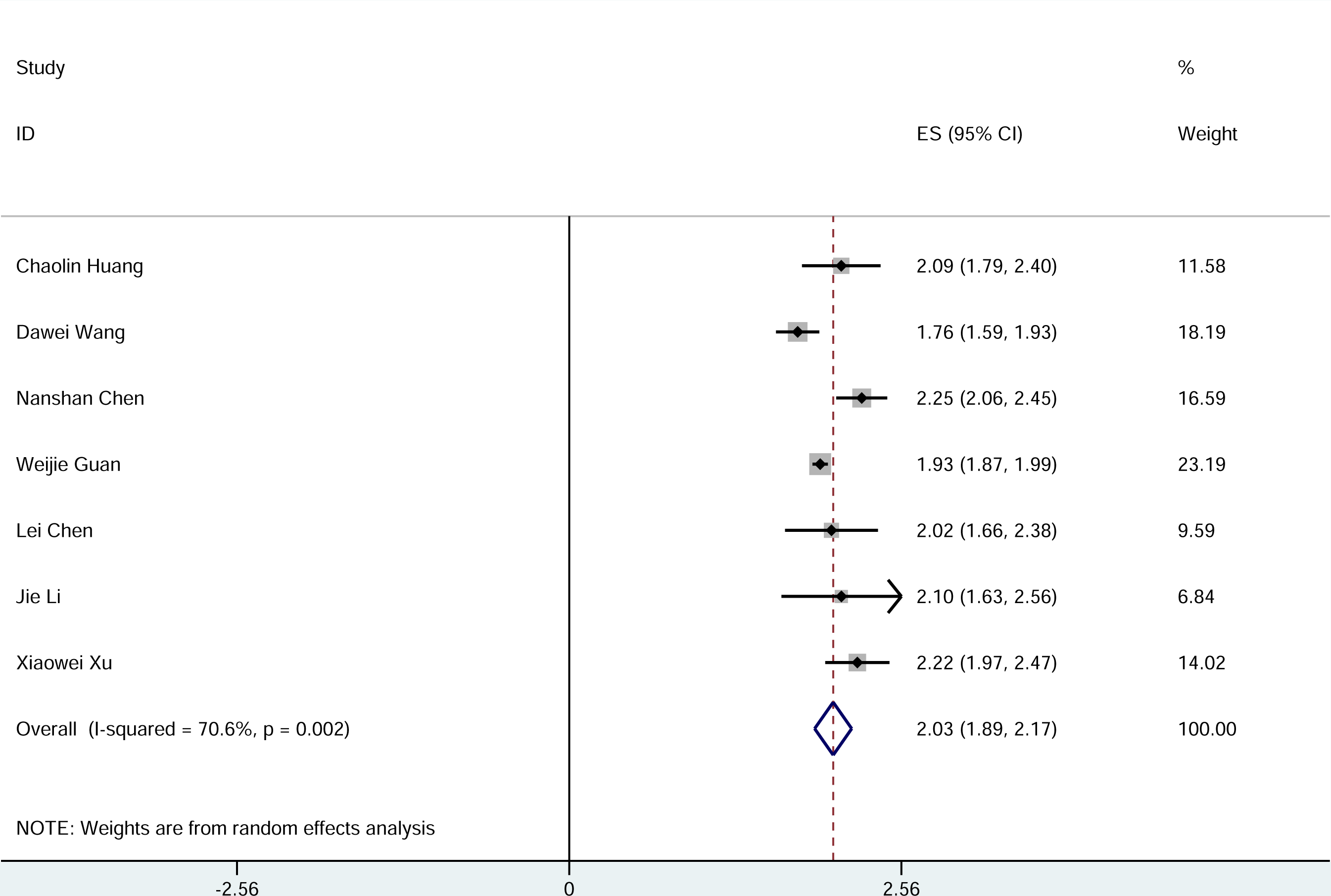
Meta-analysis of the incidence of cough.

**Figure 4.**
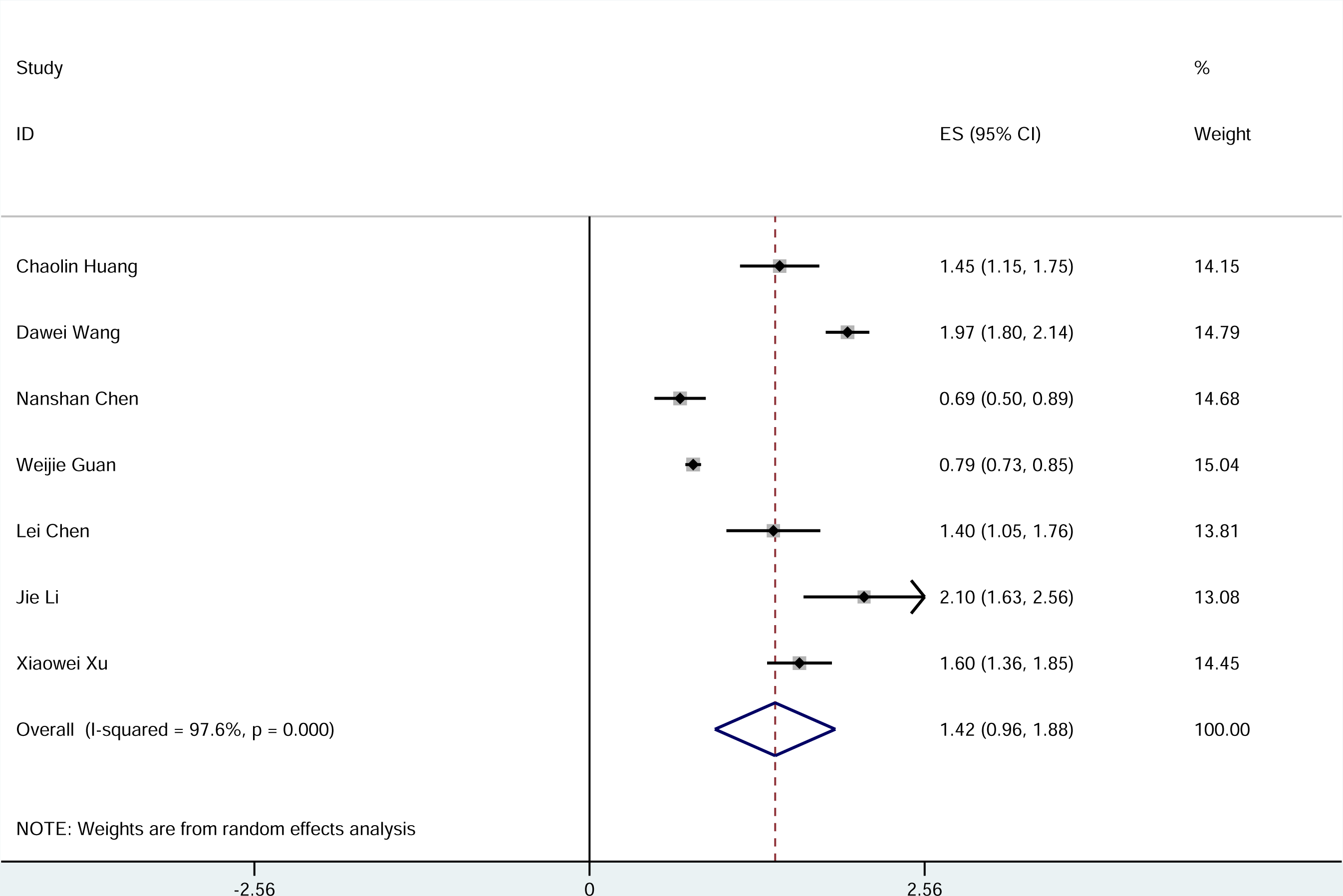
Meta-analysis of the incidence of muscle soreness or fatigue.

**Figure 5.**
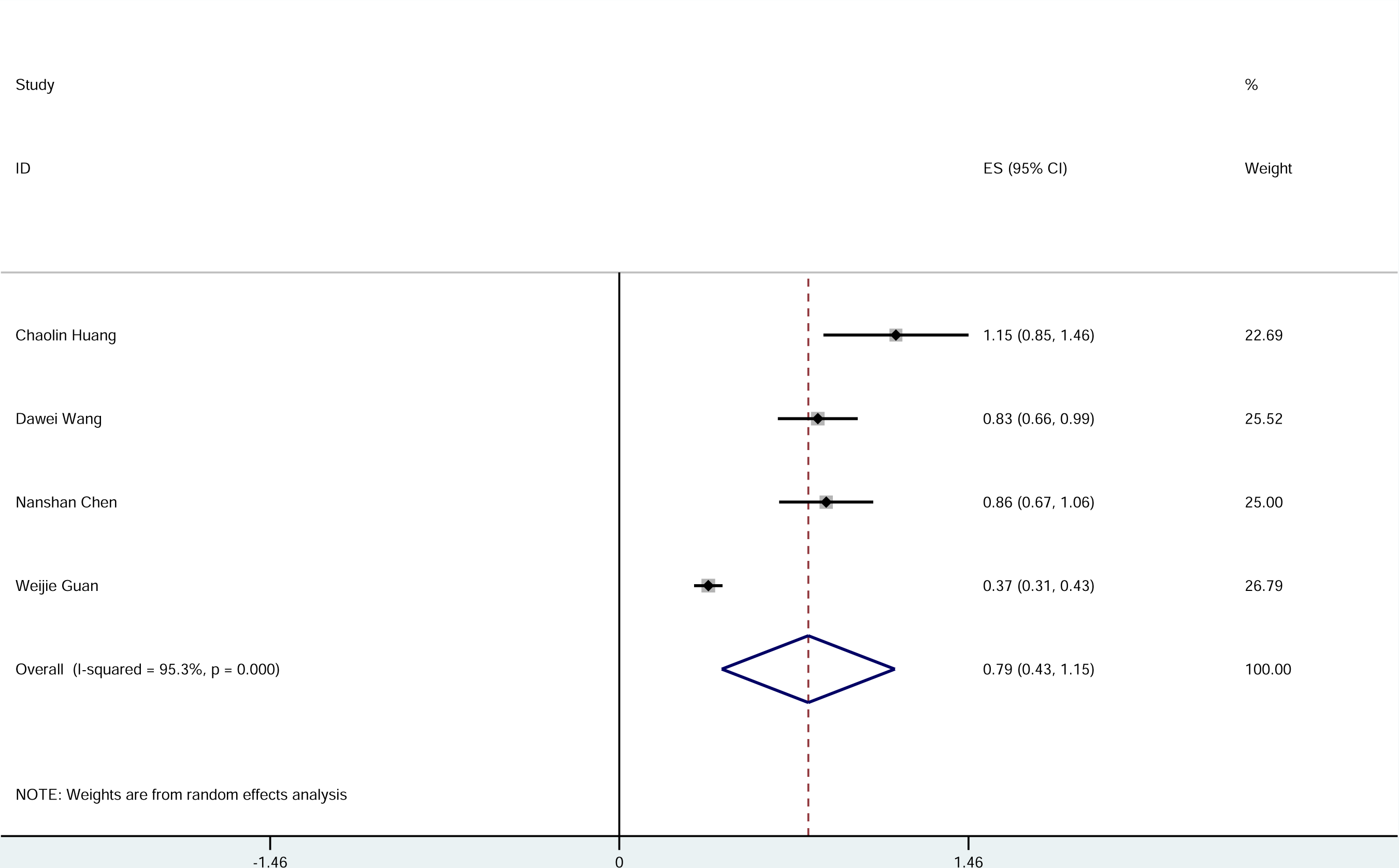
Meta-analysis of the incidence of ARDS.

**Figure 6.**
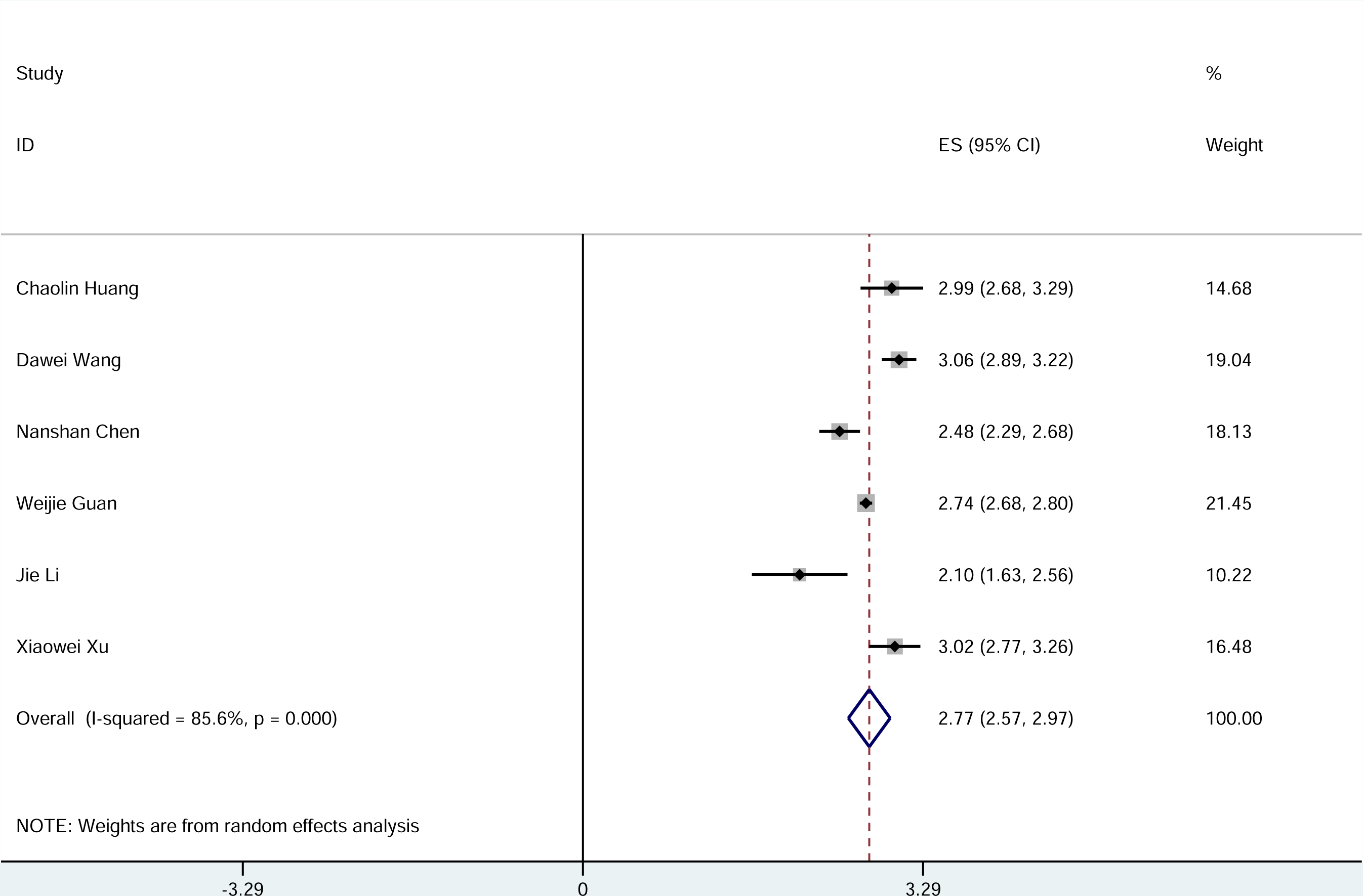
Meta-analysis of the incidence of abnormal chest CT.

**Figure 7.**
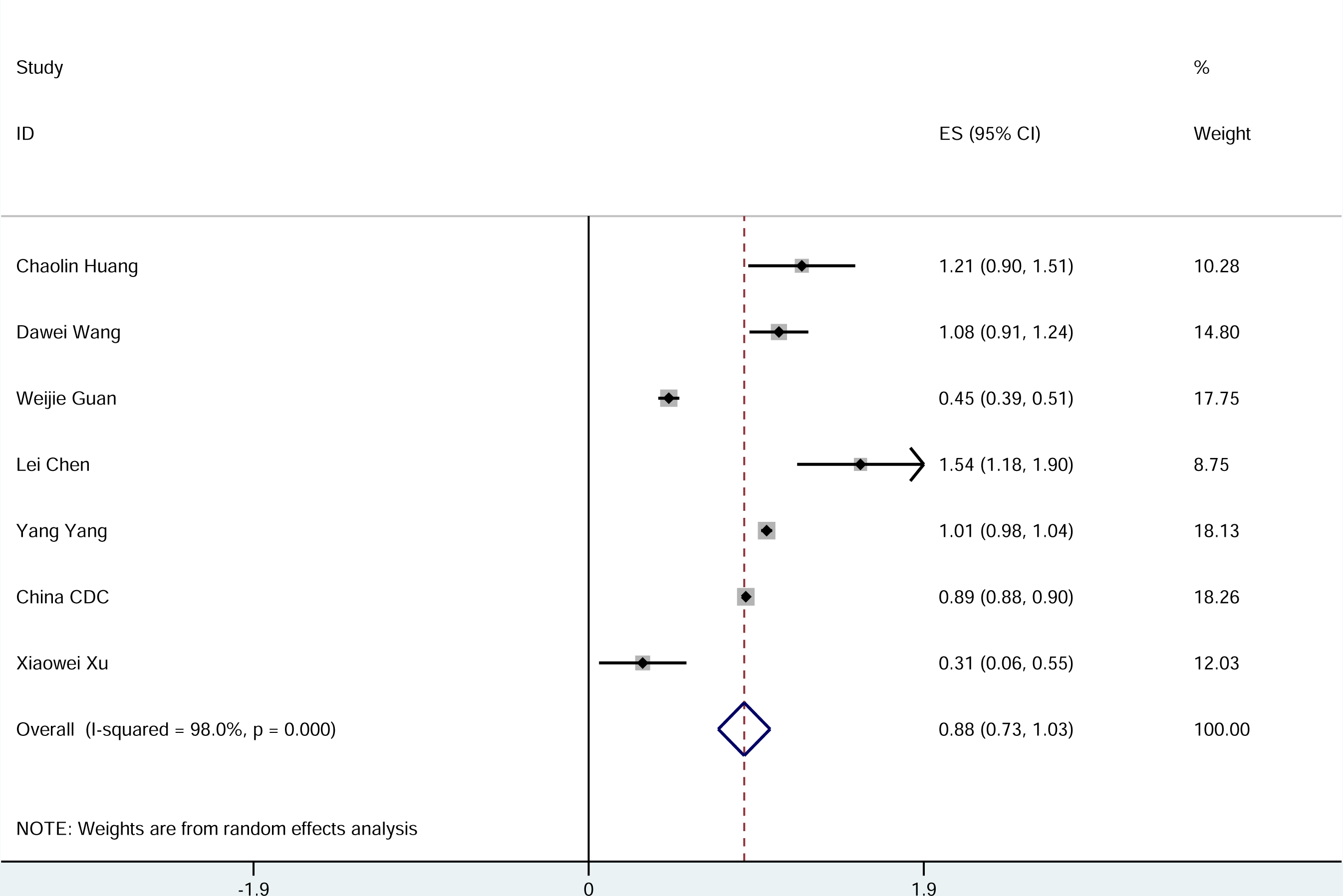
Meta-analysis of the proportion of severe cases in all infected cases.

**Figure 8.**
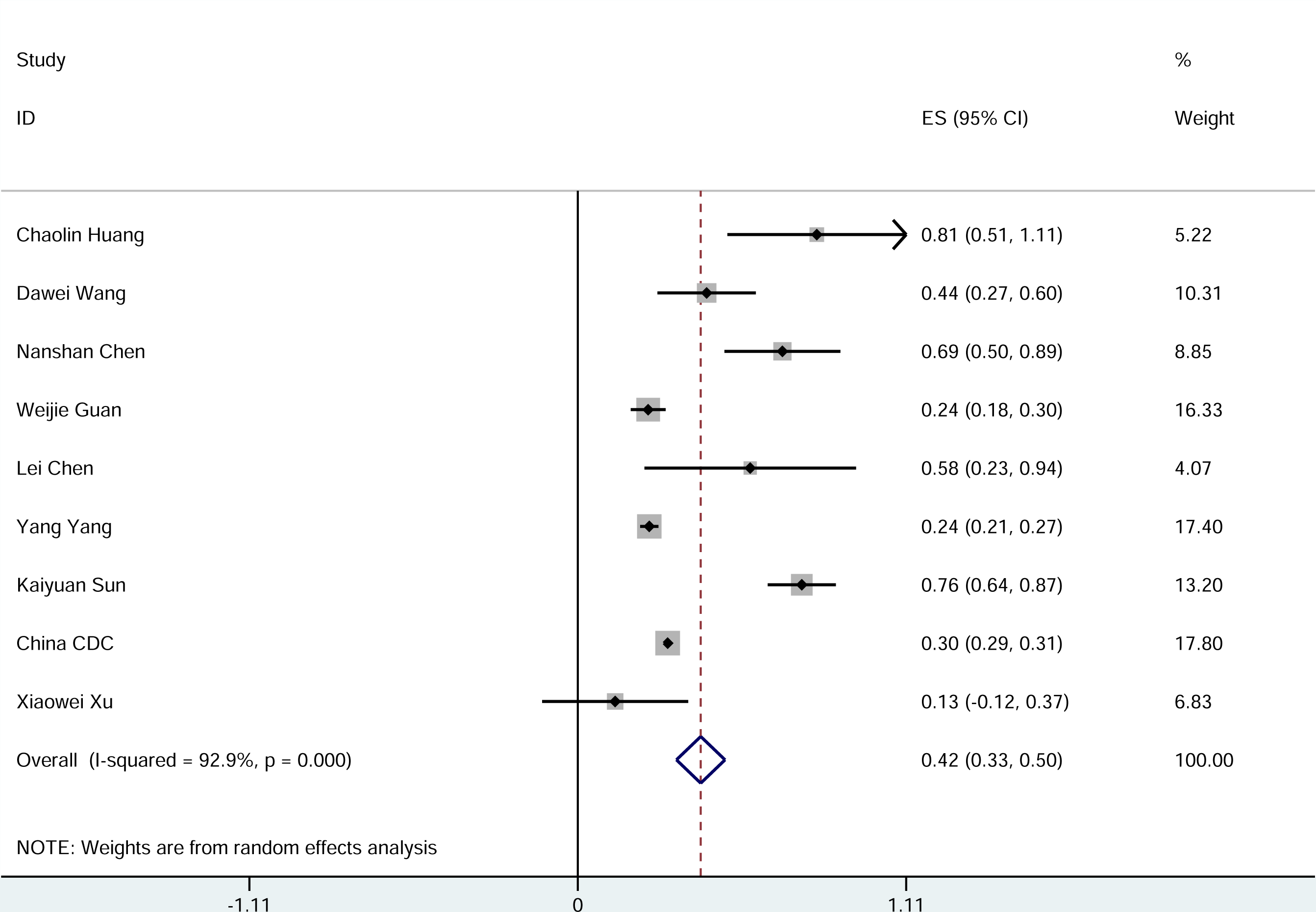
Meta-analysis of the mortality rate of patients with 2019-nCoV infection.

### 3.3 Publication bias detection

The results of Egger test in Table 3. There was publication bias in the Meta-analysis of the ARDS group(P=0.008)..

**Table 3:** Results of Egger test.

| Clinical Symptom | Fever | ARDS | Muscle Soreness or Fatigue | Cough | Abnormal Chest CT | Patient in Critical Condition | Death of Patient |
| --- | --- | --- | --- | --- | --- | --- | --- |
| P 值 | 0.866 | 0.008 | 0.090 | 0.278 | 0.908 | 0.826 | 0.258 |

## 4 Discussion

2019-nCoV is one type of coronaviruses which belongs to β-coronavirus cluster. It causes the third kind of zoonotic coronavirus disease after Severe Acute Respiratory Syndrome (SARS) and Middle East Respiratory Syndrome (MERS). Wei Ji *et al*. ^[15]^ found that 2019-nCoV seemed to be a recombinant virus between bat coronavirus and coronavirus of another unknown origin. Kangpeng Xiao *et al*. ^[16]^ confirmed that malayan pangolins were the most likely intermediate host of 2019-nCoV. The research of Domenico Benvenuto *et al*. ^[17]^ found that 2019-nCoV was only closely related to the coronavirus isolated from chrysanthemum-headed bat in 2015. Their research suggests that bats may be the reservoir host for 2019-nCoV. According to the research by Peng Zhou *et al*. ^[18]^ and Fan Wu *et al*. ^[19]^, it was found that the sequence homology between 2019-nCoV and SARS-CoV was 79.5%. The homology between 2019-nCoV and bat coronavirus at the genetic level was 96%. Therefore, very likely it can be confirmed that 2019-nCoV comes from bats.

According to the results of Meta-analysis, we find that the most common symptoms of patients with 2019-nCoV infection were fever and cough, and most of the patients had abnormal chest CT. Some patients with 2019-nCoV infection had muscle soreness or fatigue, ARDS, and other symptoms. Diarrhea, hemoptysis, headache, sore throat, shock, and other symptoms only occurred among a small number of patients. The proportion of severe cases in all infected cases was 18.1%, and the case fatality rate was 4.3%. At present, the Chinese government has announced that the case fatality rate of patients with 2019-nCoV infection in China is 3.36%, which is lower than that of another two widely contagious zoonotic coronavirus diseases, SARS and MERS. Infection mortality rates of SARS and MERS are 9.6% ^[20]^ and 35% ^[21]^, respectively.

Gaohong Sheng *et al*. ^[22]^ have shown that viral infection could increase the risk of pulmonary fibrosis. Xie *et al*. ^[23]^ found that 45% of patients showed signs of pulmonary fibrosis within one month after being infected with SARS-CoV. Hui *et al*. ^[24]^ revealed that 36% and 30% of patients infected with SARS-CoV developed pulmonary fibrosis 3 and 6 months after infection. These studies consistently suggest that pulmonary fibrosis will become one of the serious complications in patients with 2019-nCoV infection. How to prevent and reduce the occurrence of pulmonary fibrosis in patients with 2019-nCoV infection are urgent problems for medical workers in the treatment of 2019-nCoV.

Yu Zhao *et al*. ^[25]^ found that angiotensin converting enzyme 2 (ACE2) was the receptor of 2019-nCoV. In normal lung tissue, ACE2 is mainly expressed by type □ and type □ alveolar epithelial cells. It was reported that 83% of II-type alveolar cells expressed ACE2. Therefore, 2019-nCoV infection causes damages to most II type alveolar cells. In addition, the use of mechanical ventilation in the treatment of patients with 2019-nCoV infection can also aggravate the injury of alveolar cells. After alveolar cell injury, transforming growth factor-β (TGF-β) released in tissue promotes the lung repair. Virus infection often leads to excessive activation of TGF-β pathway, which leads to the occurrence of pulmonary fibrosis.

Based on the mechanism of pulmonary fibrosis after virus infection, we can formulate corresponding prevention and treatment proposals. The first is the treatment of 2019-nCoV infection. At present, a variety of drugs have been found to inhibit 2019-nCoV. Michelle L. Holshue *et al*. ^[26]^ achieved remarkable results in the treatment of patients with 2019-nCoV infection with remdesivir. This drug is currently in clinical trials. Second, it is necessary to inhibit the inflammatory reaction and reduce the exudation. On this direction, antibiotics not only have traditional antibacterial effects, but also have immunomodulatory and anti-inflammatory properties ^[27]^. Macrolide antibiotics such as clarithromycin, azithromycin, and erythromycin prevent the production of pro-inflammatory cytokines and immune mediators ^[28]^. The third is to inhibit the fibrogenic effect of TGF-β. Luo *et al*. ^[29]^ found that arsenic trioxide could inhibit the signal transduction of TGF-β and play a role in anti-fibrosis. The fourth is the rational use of ventilator to avoid unnecessary lung damages. In addition, pirfenidone and nintedanib as drugs for the treatment of idiopathic pulmonary fibrosis can also be used to prevent and treat pulmonary fibrosis in patients with 2019-nCoV infection ^[30]^.

In all, a total of 10 articles were covered in the Meta-analysis, including 50466 patients with 2019-nCoV infection. By far, it is the first Meta-analysis with the largest sample size. The quality of the literature included in this study is high, the analysis is rigorous, and the conclusions drawn by the study are highly credible. However, this Meta-analysis also has some limitations: 1) All studies included in this Meta-analysis are retrospective studies with large heterogeneity; 2) Most patients in our Meta-analysis are Chinese, and we aimed to use the conclusions of this study to predict patients in general, including other countries and races; 3) There was publication bias in the Meta-analysis of the ARDS group. The analytical conclusion of the ARDS group may be influenced by publication bias. Therefore, based on the above limitations, the conclusions of this Meta-analysis still need to be verified by more relevant studies with more careful design, more rigorous execution, and larger sample size.

## Data Availability

All data are included in the article.

## Contributors

Jianing Xi and Pengfei Sun had the idea for and designed the study and had full access to all data in the study and take responsibility for the integrity of the data and the accuracy of the data analysis. Pengfei Sun and Shuyan Qie contributed to writing of the report. Jianing Xi contributed to critical revision of the report. Shuyan Qie, Zongjan Liu and Jizhen Ren contributed to the statistical analysis. All authors contributed to data acquisition, data analysis, or data interpretation, and reviewed and approved the final version.

## Acknowledgements

The authors declare no conflict of interest in this study. We acknowledge TopEdit LLC for the linguistic editing and proofreading during the preparation of this manuscript.

